# Pharmacogenomic variant profiling in 14,490 Koreans using a population-specific genotyping array

**DOI:** 10.64898/2026.01.19.26344411

**Authors:** Sinwoo Park, Minjin Seo, Chan Hee Park, Hyun-Young Park, Young Jin Kim, Bong-Jo Kim

## Abstract

Pharmacogenomics is an essential component of precision medicine; however, most existing knowledge has been derived from populations of European ancestry, limiting the understanding of pharmacogenomic diversity in East Asian populations. In this study, we applied genotype imputation to the Korea Biobank Array v2.0 using a reference panel of 8,062 Korean whole-genome sequencing (WGS) samples and analyzed pharmacogenomic variants and phenotypes in 14,490 Korean individuals. To assess the accuracy of imputation-based variant detection, we compared imputed genotypes with matched WGS data from 735 individuals and with genotypes obtained from the commercial PangenomiX Plus Array Kit for an additional 137 individuals, demonstrating high concordance. When extended to the full cohort, all individuals were found to carry at least one pharmacogenomic variant, with high frequencies observed in key pharmacogenes including *CYP2C19*, *SLCO1B1*, *CYP3A5*, and *VKORC1*. Phenotype distributions were broadly consistent with previous WGS-based studies in East Asians but showed notable differences compared with European populations. Overall, this population-specific, large-scale analysis provides a comprehensive pharmacogenomic landscape in Koreans and highlights the importance of ancestry-tailored data for equitable precision medicine.

## Introduction

One of the central goals in biomedical science is the realization of precision medicine tailored to individual patients^1^. Optimizing drug therapy based on genomic information is a key component of precision medicine. Although most drugs are prescribed at standardized doses, interindividual genetic differences can profoundly affect both therapeutic efficacy and the risk of adverse reactions^2^. Indeed, a substantial proportion of individuals are estimated to carry at least one variant that alters drug metabolism or response^3,4^. Consequently, pharmacogenomics (PGx), which examines how genetic variation influences drug response, has become a cornerstone of efforts to implement personalized medicine.

After administration, drugs undergo absorption, distribution, metabolism, and excretion (ADME)^5^. These processes are controlled by multiple genes, including *CYP2C19*, *VKORC1*, *CYP3A5*, and *SLCO1B1*, which regulate drug transport, activation, and clearance^6–9^. Metabolism—largely mediated by the cytochrome P450 (CYP) family in the liver—is particularly critical^10^. Polymorphisms in *CYP2C9* and *CYP2C19* are well known to alter enzymatic activity, leading to differences in drug exposure and toxicity^11^. *CYP3A5* is polymorphically expressed, and the nonfunctional *3 allele reduces the clearance of drugs such as tacrolimus and simvastatin^8^. Warfarin response is strongly influenced by variants in *VKORC1*, which encodes the molecular target of this anticoagulant^7^. Similarly, reduced-function alleles in *SLCO1B1* impair statin transport, increasing systemic drug levels and the risk of myopathy^9^. These examples underscore the clinical importance of PGx variation across diverse therapeutic areas.

The translation of such findings into clinical practice has been facilitated by dedicated resources. PharmGKB curates information on gene–drug relationships, genotype–phenotype associations, and dosing guidelines^12^, whereas PharmVar standardizes star allele definitions for clinically important pharmacogenes^13^. Implementation guidelines from the Clinical Pharmacogenetics Implementation Consortium (CPIC) and the Dutch Pharmacogenetics Working Group provide evidence-based recommendations to guide clinicians^14^. Collectively, these resources establish a framework for linking genotypes to clinical decision-making and have laid the groundwork for precision prescribing.

Despite this progress, most PGx resources and large-scale studies have been developed using data from individuals of European ancestry. For instance, the All of Us program has enrolled approximately 245,000 participants and the UK Biobank over 200,000, both primarily of European descent^15–18^. In contrast, data from East Asia remain relatively scarce. The Taiwan Biobank includes 172,854 individuals—far fewer than major European cohorts—and only one Korean study has been conducted to date, involving 396 participants^19–21^. Because gene–drug interaction studies depend on large, population-scale cohorts, the limited representation of East Asian populations poses a significant limitation in pharmacogenomic research.

Recognizing this gap, we conducted a comprehensive pharmacogenomic analysis of 14,490 Korean individuals using the Korea Biobank Array v2.0 (KBAv2.0), which includes 1.66 million variants, approximately 282,000 of which are pharmacogenetic or clinically actionable^22^. By characterizing the distribution and functional implications of key pharmacogenetic variants, this study establishes a population-specific reference for pharmacogenomic implementation in Koreans and advances the understanding of pharmacogenomic diversity in East Asian populations. Furthermore, we systematically compared haplotype and phenotype frequencies of key pharmacogenes across multiple ancestral groups, including East Asian and European populations, to identify population-specific patterns that may inform ancestry-guided precision prescribing.

## Materials and Methods

### Preprocessing and quality control of KBAv2.0

We utilized genotyping data from 14,529 participants in the Korean Genome and Epidemiology Study (KoGES). Genotype calling for the KBAv2.0 array was performed using Analysis Power Tools (APT, v2.11.8) with the Axiom_KORv1_1.r1 and Axiom_KBAbetaB_r2_3 library files, applying the –do-rare-het-adjustment option. Single-nucleotide polymorphism (SNP)–specific high-quality variants were selected using SNPolisher^23^, followed by an additional round of genotype calling with SNP-specific priors and advanced normalization in APT to further refine the dataset.

After genotype calling, sample-level quality control was conducted. Samples with a missing rate > 0.03 or heterozygosity outside the range of 15–20 were excluded. Outlier detection based on population structure was performed using FlashPCA^24^. Additionally, samples identified as duplicates through relationship inference using KING (v2.3.2)^25^ and those with sex discrepancies were removed, resulting in a final dataset of 14,490 samples.

From the 14,490 samples, variants with a call rate < 95%, Hardy–Weinberg equilibrium (HWE) p-value <1 × 10⁻⁶, and fixation index (F_st_) > 0.025, compared to variant frequencies in 8,062 whole genome sequencing data from the Korean Imputation Service^26^ (https://coda.nih.go.kr/usab/kis/intro.do), were removed. When restricted to autosomal variants with a minor allele frequency (MAF) ≥ 0.01, a total of 875,789 variants were retained from initial 1.66 million variants. Phasing and imputation of the preprocessed KBAv2.0 data were performed using a reference panel of 8,062 Korean WGS samples^26^. Phasing was conducted with Eagle (v2.4.1)^27^, followed by reference-based imputation using Minimac4 (v1.0.2)^28^. After imputation, a total of 47,862,506 variants were retained, constituting the final dataset for downstream pharmacogenomic analyses.

### Pharmacogenomic variant calling and genotypic analysis using PharmCAT

Pharmacogenomic variant calling was performed using PharmCAT (v3.0.0)^29^, which requires genotyping data in the GRCh38 reference build. Accordingly, GRCh37-based genotyping data were converted to GRCh38 using GATK^30^ LiftoverVcf with the --RECOVER_SWAPPED_REF_ALT option. Genotyping variants from KBAv2.0 were then filtered to retain only those corresponding to allele-defining positions specified by PharmCAT. The multi-sample VCF was divided into individual files and preprocessed using the PharmCAT VCF Preprocessor to normalize and format the data, ensuring that all allele-defining loci were included. PGx variant calling was conducted in researcher mode using PharmCAT, which produces genotype-level outputs without clinical interpretation and is intended solely for research purposes. Following PGx variant calling, the resulting PharmCAT reports were post-processed using R (v4.4.3) and Python (v3.11.1) to generate summary tables and visualizations. In the analysis, the *G6PD* gene was excluded because the WGS reference panel used for imputation lacked sex chromosome data, preventing reliable analysis despite its presence in KBAv2.0. The *CYP2D6*^31^ and *HLA*^32^ genes were also excluded because their complex genomic architectures require specialized genotyping approaches. Consequently, a total of 18 genes including *CYP2C19*, *CYP2C9*, and *VKORC1*, were retained for analysis.

Genotyping data were obtained from the KoGES and are available to qualified researchers upon request from the National Biobank of Korea (NBK, Cheongju, Republic of Korea; http://biobank.nih.go.kr).

### Preprocessing of datasets for KBAv2.0 performance evaluation

We used two independent datasets to evaluate PGx variant detection with KBAv2.0: the PangenomiX Plus Array (PangenomiX Plus; Thermo Fisher Scientific, Waltham, MA, USA)—a chip specifically designed for pharmacogenomic variant detection containing approximately 850,000 markers, including variants cataloged in CPIC and PharmGKB—and high-coverage WGS data from 8,062 Koreans. Preprocessing and quality control (QC) were performed separately for each dataset before cross-platform comparison.

Genotyping data generated from the PangenomiX Plus were processed according to established procedures. Raw intensity (CEL) files were analyzed using APT (v2.11.8) with the Axiom_PangenomiX.r1 library, and genotype calling with SNP–specific quality assessment was performed using SNPolisher. High-quality SNPs were selected based on these classifications. Sample-level quality control was also performed following the same procedures as for KBAv2.0, except that samples with heterozygosity outside the range of 23–24.5 were excluded. Among the initial 1,056 participants, no duplicate individuals were identified, and a total of 27 samples were excluded, resulting in 1,029 individuals for the final analysis. Additionally, SNP-level quality control was conducted following the procedures applied to KBAv2.0. After selecting common variants with a MAF ≥ 0.01, a total of 443,203 variants were retained from the initial 868,387 variants. Following genotyping and QC, 1,029 samples and 443,203 variants were included in downstream analyses. Subsequent preprocessing mirrored the KBAv2.0 pipeline, incorporating guided phasing with Eagle (v2.4.1) and reference-based imputation with Minimac4 (v1.0.2) using the same Korean WGS reference panel. After imputation, 47,862,502 variants were identified across the 1,029 samples, yielding a comprehensive genome-wide dataset for subsequent pharmacogenomic analyses.

In the case of WGS data^26^, we followed the preprocessing procedures established for constructing the Korean Reference Genome panel. Briefly, raw sequencing reads were quality-controlled using Trimmomatic (v0.39), aligned to the GRCh37 reference genome, and polymerase chain reaction duplicates were removed^33^. Variant discovery was performed using the DRAGEN™ platform (Illumina Inc., San Diego, CA, USA), and raw variants were further refined using the GATK Variant Quality Score Recalibration process. After applying both sample- and variant-level quality control filters, 20,220,176 high-quality variants from 8,062 WGS samples were retained for subsequent analyses.

### Performance evaluation of KBAv2.0

The performance of KBAv2.0-derived PGx variant detection was assessed through cross-platform comparisons. To ensure robust validation, we leveraged a large cohort comprising 735 individuals genotyped by both KBAv2.0 and WGS, along with 137 individuals genotyped by both KBAv2.0 and PangenomiX Plus. For each PharmCAT-defined site, the presence of alternate alleles was compared across platforms, and performance was quantified using precision, recall, and the F1 score.

To further validate the consistency of KBAv2.0-derived PGx calls, we additionally referenced PGx haplotype and phenotype data from a previously published WGS-based study involving 396 Korean individuals^20^. Seven genes—*CACNA1S*, *CFTR*, *CYP4F2*, *IFNL3*, *RYR1*, *VKORC1*, and *CYP3A4*—were excluded because their nomenclature classifications differed between the previous study and the KBAv2.0-based results.

### Comparison of population-level pharmacogenomic phenotypes

We compared pharmacogenomic characteristics observed in the KBAv2.0 dataset with those reported in other ancestral populations using haplotype and phenotype data from several large-scale genomic resources, including the All of Us (AoU) Research Program, the UK Biobank (UKBB), the 1000 Genomes Project (1000G), and the Taiwan Biobank (TWB). For AoU and UKBB, data were obtained from PharmGKB^34^. Whereas TWB^21^ and 1000G^35^ data were extracted from previously published studies. To enable accurate ancestry-specific comparisons, the UKBB subgroup combining individuals of South Asian and Central Asian ancestry was excluded. Additionally, participants from any cohort who were not assigned to a specific ancestry group were excluded. In the TWB dataset, *TPMT* and *VKORC1* were excluded because their nomenclature classifications from reported phenotypes did not align with those observed in this study and in previously published studies.

### Statistical analysis for population-level comparison

We compared the number of pharmacogenomic variants between the Korean population and the 1000G cohort using Welch’s t-test^36^. To minimize the potential influence of differences in sample size, we additionally performed bootstrapping with 10,000 replicates. Furthermore, to visualize and compare haplotype and pharmacogenomic phenotype distributions across ancestral groups in the 1000G cohort, we performed dimensionality reduction using Uniform Manifold Approximation and Projection, implemented in the uwot package in R (v4.3.3).

Statistical comparisons of pharmacogenomic phenotype distributions between Koreans and each comparator population were conducted using either chi-square tests or Fisher’s exact tests. The chi-square test and Fisher’s exact test were performed in R. The chi-square test was applied when all expected counts were ≥ 5; otherwise, Fisher’s exact test was used. To account for differences in sample size across populations and cohorts, effect sizes were calculated using Cramér’s V, implemented in the DescTools package (v0.99.6) in R^37,38^. Phenotype categories labeled as “Indeterminate” or “Others,” which lacked clear classification, were excluded from statistical testing.

## Results

### Benchmarking the performance of KBAv2.0 against sequencing and array-based platforms

Genotyping arrays directly measure only a predefined set of variants and rely on imputation to infer additional variants, which may affect accuracy. To evaluate the accuracy and reliability of KBAv2.0, we benchmarked its performance against both sequencing- and array-based platforms using matched cohorts of considerable size—735 individuals genotyped by both KBAv2.0 and WGS, and 137 individuals genotyped by both KBAv2.0 and PangenomiX Plus.

Among the 759 pharmacogenomic sites analyzed, 71 were consistently identified as polymorphic in both KBAv2.0 and WGS, with no discordant genotypes (F1 = 1.0; Figure 1a and Supplementary Table S1). This finding indicates complete concordance in genotype detection at the site level between KBAv2.0 and WGS. Across 735 individuals, carrier frequencies for these polymorphic sites were nearly identical (Spearman’s r = 0.999; Figure 1b). Minor discrepancies in carrier frequencies were observed at two loci: rs2279343 (*CYP2B6*), with frequencies of 33.197% in KBAv2.0 and 34.558% in WGS, and rs4020346 (*CYP4F2*), with frequencies of 1.088% in KBAv2.0 and 0.816% in WGS. These small differences had minimal impact on downstream interpretation, as pharmacogenomic phenotypes remained highly consistent (r = 0.999), with discrepancies confined to *CYP2B6* and *CYP4F2*, while all other genes showed identical phenotype assignments (Figure 1c).

**Figure 1.**
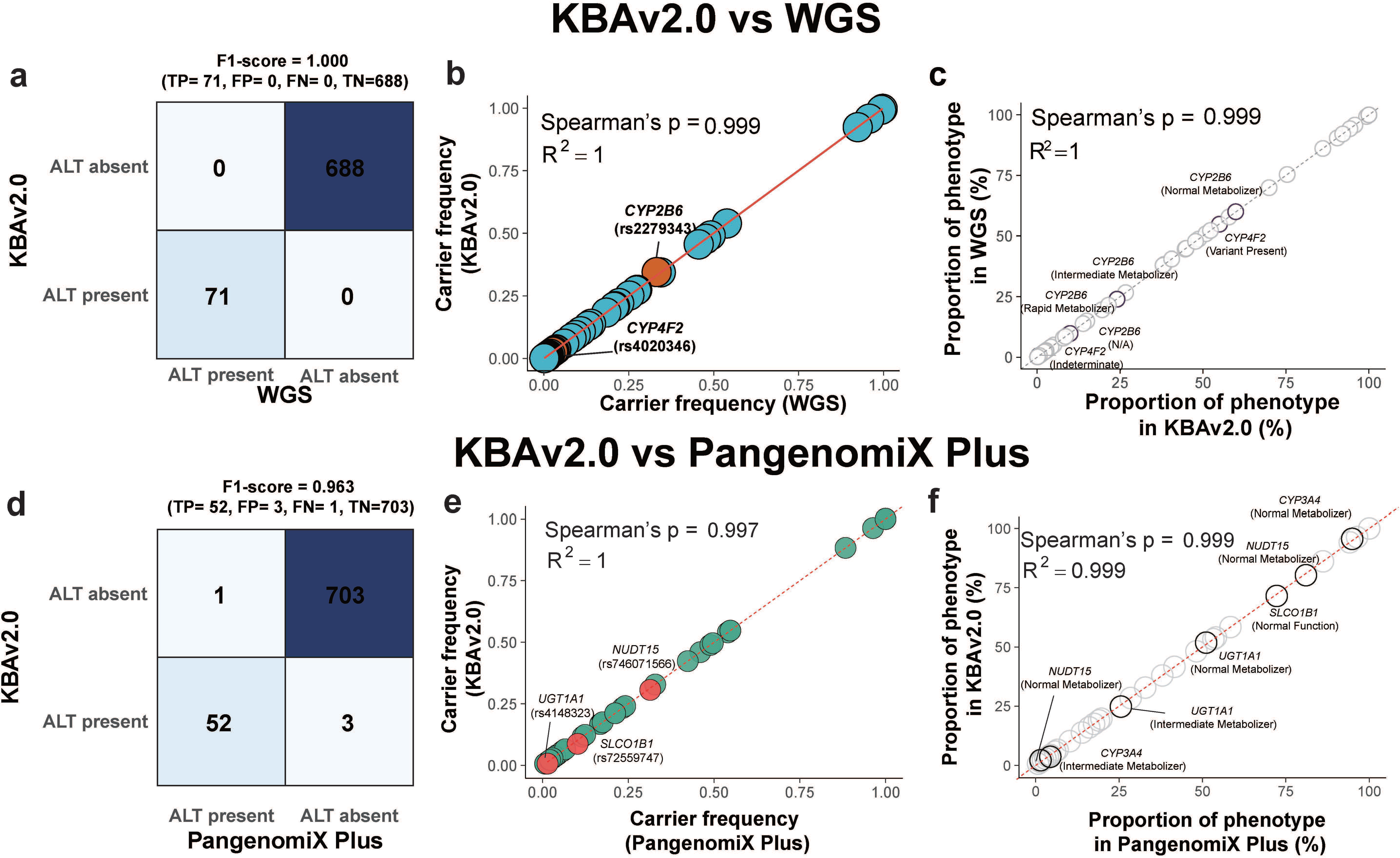
Evaluation of imputation-based pharmacogenomic variant calling in the KBAv2.0 array using WGS and PangenomiX Plus. **(a)** Comparison of pharmacogenomic variant detection between WGS and KBAv2.0. “ALT present” indicates the number of variant sites carrying alternate alleles, whereas “ALT absent” represents homozygous reference sites with no detected variant. **(b)** Comparison of carrier frequencies for 71 pharmacogenomic variants commonly identified in both WGS and KBAv2.0. Red dots denote variants showing frequency differences between the two platforms; the red line indicates the Y = X line. **(c)** Comparison of pharmacogenomic phenotype frequencies derived from WGS and KBAv2.0. Dotted points mark gene–phenotype pairs showing differences in phenotype frequency between platforms. **(d)** Comparison of pharmacogenomic variant detection between PangenomiX Plus and KBAv2.0. As in panel A, “ALT present” and “ALT absent” indicate variant-containing and homozygous reference sites, respectively. **(e)** Comparison of carrier frequencies for 52 pharmacogenomic variants commonly identified in both PangenomiX Plus and KBAv2.0. Red dots highlight variants with differing frequencies between the two platforms; the red line indicates the Y = X line. **(f)** Comparison of pharmacogenomic phenotype frequencies identified by PangenomiX Plus and KBAv2.0. Bold points represent gene–phenotype pairs exhibiting different phenotype frequencies between platforms; the red line denotes the Y = X line.

To further evaluate the reliability of KBAv2.0, we compared it with the PangenomiX Plus across 759 pharmacogenomic sites. Genotype-level concordance was high (F1 = 0.963; Figure 1d and Supplementary Table S2). KBAv2.0 detected alternate alleles at 55 sites, whereas PangenomiX Plus detected 53 sites, 52 of which were shared between the two platforms. For these 52 polymorphic sites, carrier frequencies among 137 individuals were highly correlated (Spearman’s r = 0.997; Figure 1e). Minor discrepancies were observed at three loci, rs746071566 (*NUDT15*), rs72559747 (*SLCO1B1*), and rs4148323 (*UGT1A1*), while the remaining 49 sites showed identical frequencies. Importantly, pharmacogenomic phenotypes were also highly consistent (r = 0.999), with minor differences confined to a few genes such as *UGT1A1*, *CYP3A4*, and *SLCO1B1* (Figure 1f).

Overall, these results demonstrate that the imputation-based application of KBAv2.0 achieves performance comparable not only to sequencing-based platforms but also to commercial pharmacogenomic arrays.

### Genetic variability of pharmacogenomic loci in KBAv2.0 from 14,490 Koreans

Building on the high concordance of KBAv2.0 with sequencing and commercial pharmacogenomic array platforms, we extended the analysis to 14,490 Korean individuals. Among the 759 pharmacogenomic loci, 663 (87.352%) were monomorphic, with all individuals carrying the homozygous reference genotype and showing no observed variation at these sites. In contrast, 96 loci (12.648%) were polymorphic (Supplementary Table S3 and Figure 2a). Compared with analyses of the smaller matched WGS and PangenomiX Plus datasets, the larger cohort analysis identified additional pharmacogenomic polymorphic variants, suggesting that the increased sample size enabled the detection of Korean-specific variants. At the individual level, all participants carried at least one alternate allele at the pharmacogenomic loci. On average, each individual harbored 10.163 alternate alleles across the 96 polymorphic sites, with counts ranging from 3 to 21 (Figure 2b). We also examined sample-wise carrier rates across the 96 polymorphic PGx loci (Figure 2c). On average, each locus had a carrier rate of 10.586%. Among these, loci corresponding to rs3758581 (*CYP2C19*), rs9923231 (*VKORC1*), rs776746 (*CYP3A5*), and rs2306283 (*SLCO1B1*) displayed particularly high carrier rates, with an average of 96.64% across individuals. To further explore gene-level variation (Figure 2d and Supplementary Table S4), the overall average carrier rate across all genes was 42.976%. In contrast, four genes—*CYP2C19* (99.779%), *SLCO1B1* (99.358%), *CYP3A5* (94.783%), and *VKORC1* (93.637%)—showed markedly higher carrier rates. Conversely, very low carrier rates were observed for *CACNA1S* and *CFTR*, with only 0.03% and 0.01% of individuals carrying variants in these genes, respectively.

**Figure 2.**
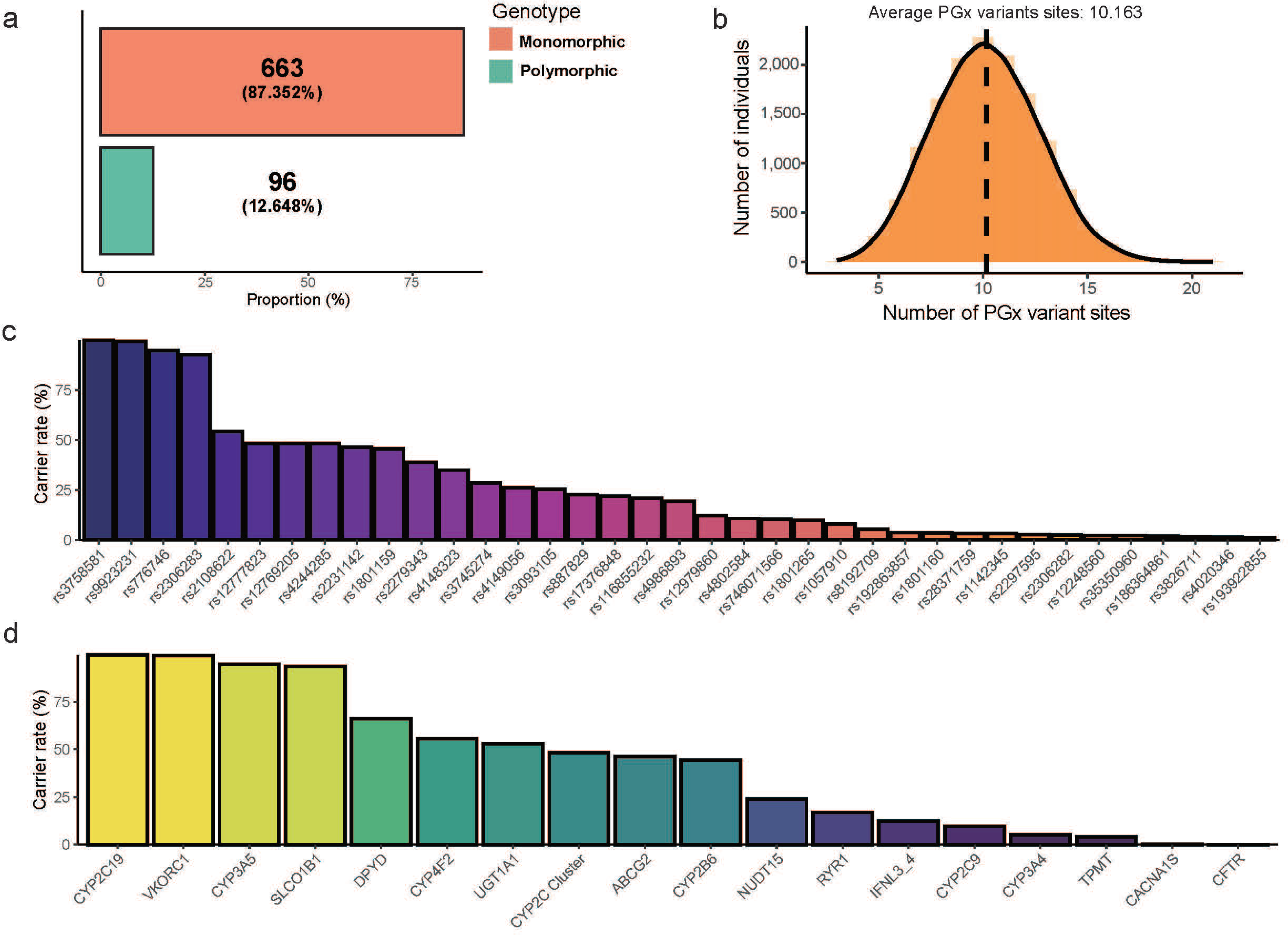
Pharmacogenomic variant profiles in 14,490 Korean individuals. **(a)** Proportion of monomorphic and polymorphic pharmacogenomic variants identified in 14,490 individuals. **(b)** Distribution of the number of pharmacogenomic variant sites carried per individual; the black dashed line indicates the average number of variants per person. **(c)** Carrier frequencies of 37 pharmacogenomic variants with a frequency greater than 1%. **(d)** Gene-wise carrier rates of pharmacogenomic variants observed across 14,490 individuals.

These findings highlight substantial variability in variant frequency across pharmacogenes and reveal that every individual in the Korean population harbored at least one pharmacogenomic variant. Notably, for several key pharmacogenes such as *CYP2C19*, *SLCO1B1*, *VKORC1*, and *CYP3A5*, nearly all individuals (>94%) carried at least one variant, indicating that genetic variation in these loci is essentially ubiquitous in the Korean population.

### Predicted pharmacogenomic phenotypes in the Korean population

We examined allelic combinations, including star alleles and gene-specific haplotypes, as these form the basis for pharmacogenomic phenotype assignment (Supplementary Figure S1 and Supplementary Table S5). Among the 18 pharmacogenes, *CYP2C19* *1 and *2 alleles accounted for 55.438% and 27.943% of total alleles, respectively. For *VKORC1* (rs9923231), 26,806 of 28,980 observed haplotypes (92.498%) harbored at least one variant. Similarly, for *CYP3A5*, the *3 haplotype was highly prevalent, accounting for 22,203 haplotypes (76.615%). In *DPYD*, *RYR1*, *SLCO1B1*, and *CYP2B6*, we observed several haplotypes containing previously unannotated variant combinations, suggesting the existence of potentially novel functional alleles in the Korean population.

To translate allelic variation into clinically relevant insights, we investigated pharmacogenomic phenotypes across the 18 genes in the Korean cohort (Supplementary Table S6). For *VKORC1*, 14,397 individuals (99.358%) carried a variant-associated phenotype, underscoring its broad influence on warfarin response (Figure 3a). In *CYP2C19*, 47.412% of individuals were classified as intermediate metabolizers (Figure 3b), suggesting that standard clopidogrel dosing may be suboptimal for a large proportion of the population. *CYP3A5* showed a striking distribution, with 58.447% classified as poor and 36.335% as intermediate metabolizers (Figure 3c), indicating a substantially reduced capacity for tacrolimus metabolism. At *SLCO1B1*, most individuals exhibited normal transporter activity (70.669%), while 23.458% exhibited decreased function (Supplementary Figure S2a), consistent with an increased risk of statin-associated adverse effects. More than half of the population (55.362%) carried a *CYP4F2* variant allele (Supplementary Figure S2b), again suggesting a potential need for genotype-guided warfarin dose adjustment. Within the *CYP2C Cluster*, the distribution of variant carriers and non-carriers was approximately balanced, with nearly equal frequencies of variant-present and variant-absent individuals (Supplementary Figure S2c). For *ABCG2*, 39.041% of individuals carried decreased-function variants (Supplementary Figure S2d), suggesting a potential impact on response to allopurinol and certain chemotherapeutic agents. In contrast, 10 genes—including *CYP2B6*, *CYP3A4*, and *CYP2C9*—showed minimal evidence of phenotype-altering variants (Figure 3e).

**Figure 3.**
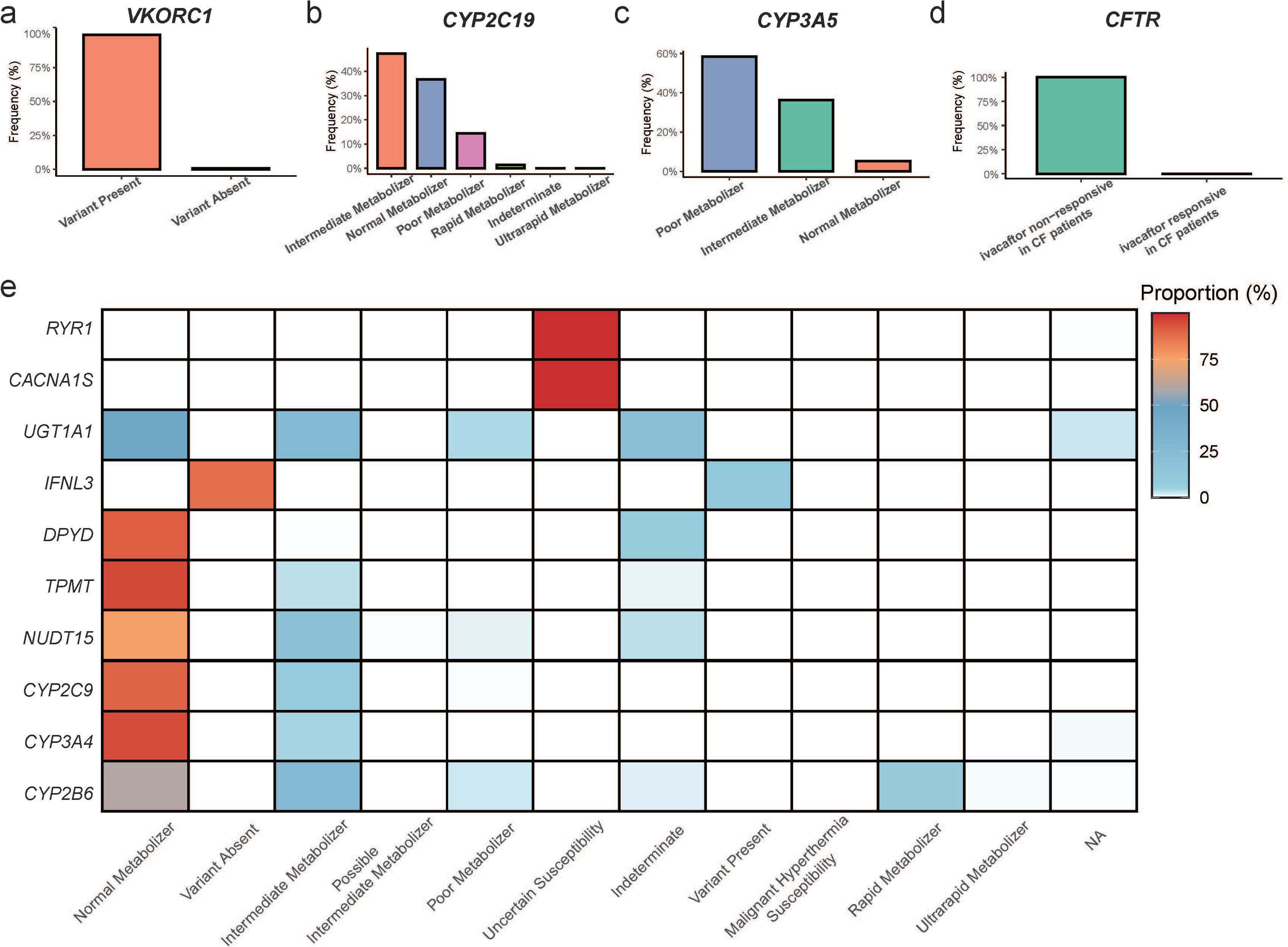
Distribution of pharmacogenomic phenotypes in 14,490 Korean individuals. **(a)** ‒ **(d)** Proportions of pharmacogenomic phenotypes for the *VKORC1*, *CYP2C19*, *CYP3A5*, and *CFTR* genes among 14,490 individuals. **(e)** Heatmap showing the distribution of pharmacogenomic phenotypes across ten genes, including *RYR1*, *CACNA1S*, and *UGT1A1*; color intensity ranges from blue to red, with red indicating higher carrier frequencies.

### Population-level comparison of pharmacogenomic variant and haplotype composition

Genetic variation differs substantially among populations due to factors such as evolutionary history, demographic structure, and migration, which collectively influence traits including disease susceptibility and drug response. As pharmacogenomic variants constitute part of this broader genomic diversity, their distributions are also expected to vary by ancestry. To investigate this, we analyzed pharmacogenomic variants and haplotypes in a large Korean cohort and compared them with multiple global populations.

Because most pharmacogenomic variants have been identified in individuals of European ancestry, we assessed whether Europeans carry a higher number of known variants than East Asians, including Koreans. Consistent with this expectation, East Asians harbored an average of 10.152 variants per individual, compared with 11.607 in Europeans (Figure 4a). Koreans specifically carried 10.163 variants, significantly fewer than Europeans (Figure 4b). Bootstrap analysis confirmed the robustness of this difference (Supplementary Figure S3), highlighting that the existing catalog of pharmacogenomic variants is likely disproportionately shaped by discoveries in European cohorts.

**Figure 4.**
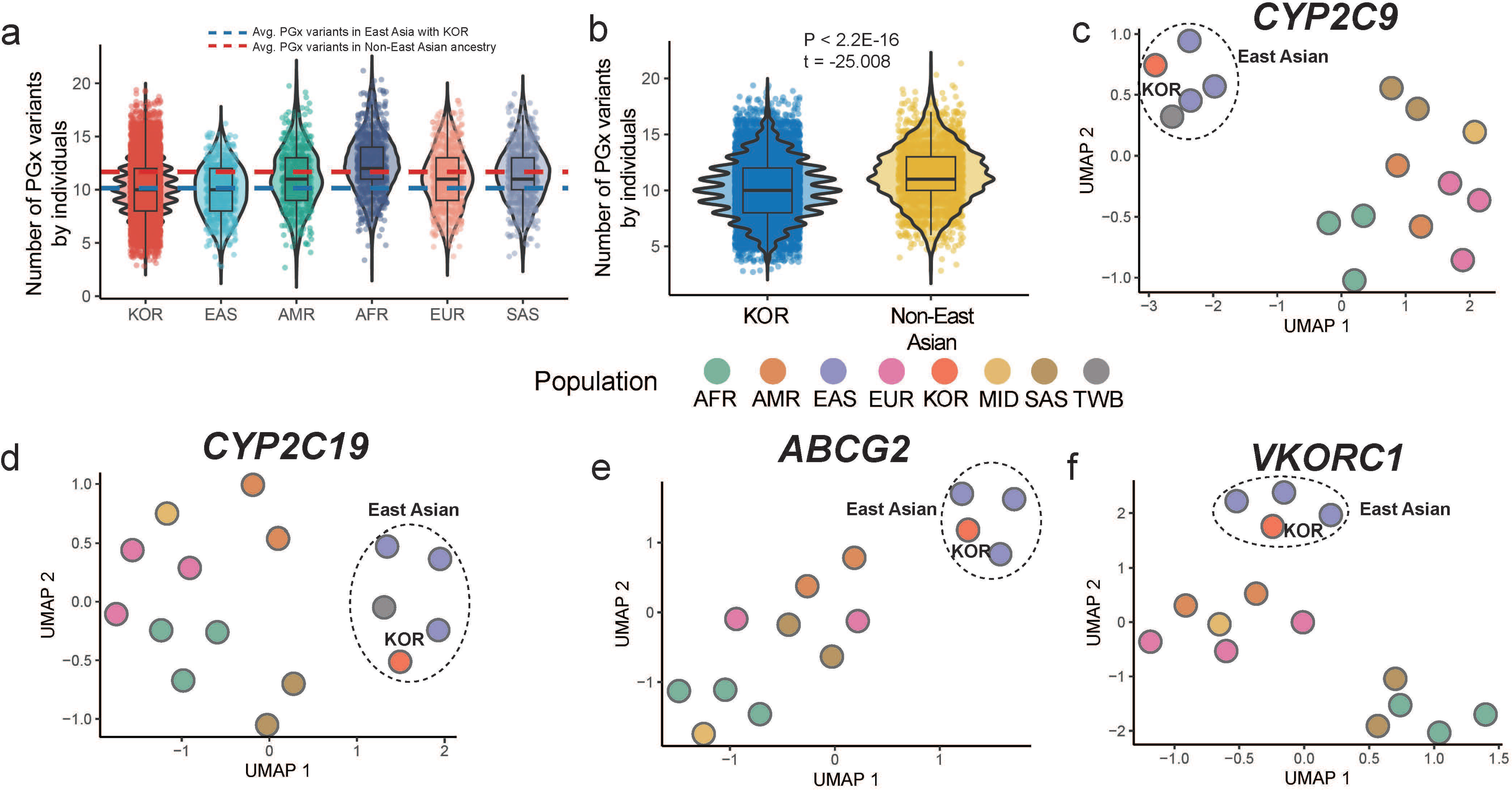
Comparison of pharmacogenomic variant and haplotype distributions across populations. **(a)** Comparison of population-level pharmacogenomic variant sites between the 1000G and KBAv2.0 data. The red dashed line indicates the average number of pharmacogenomic variants observed in non–East Asian populations, whereas the blue line represents the number observed in East Asian populations, including Koreans. **(b)** Comparison of the number of pharmacogenomic variants identified in the Korean population versus non–East Asian populations. P-values and t-statistics were calculated using Welch’s t-test. **(c)** ‒ **(f)** UMAP visualization of pharmacogenomic haplotype distributions for *CYP2C9*, *CYP2C19*, *ABCG2*, and *VKORC1* genes across different ancestry groups.

Based on the identified pharmacogenomic variants, we further examined the presence of Korean-specific haplotypes among those observed in the Korean population. Of the 168 haplotypes identified, 35 were not observed in other populations, representing Korean-specific haplotypes across eight genes, including *CYP2B6*, *CYP2C19*, and *CYP3A4* (Supplementary Table S7). Most of these haplotypes occurred at low frequencies (allele frequency < 1%); nevertheless, their presence underscores the unique pharmacogenomic diversity within the Korean population.

Furthermore, we compared the distribution of haplotype combinations between the Korean population and other ancestral groups. We hypothesized that Koreans would exhibit haplotype distributions similar to those of the broader East Asian group but distinct from those observed in Europeans. Consistent with this expectation, haplotype distributions in Koreans closely resembled those of the broader East Asian group (Supplementary Figure S4). Notably, genes such as *CYP2C9*, *CYP2C19*, *ABCG2*, and *VKORC1* exhibited strong similarity between Koreans and East Asians (Figure 4c–Figure 4f). In contrast, genes such as *CACNA1S*, *CFTR*, *DPYD*, and *RYR1* showed minimal differences in haplotype distributions across populations, suggesting a more conserved haplotype structure for these loci irrespective of ancestry.

### Population-level comparison of pharmacogenomic phenotypes

Pharmacogenomic phenotype distributions can vary even within the same ancestry due to differences in study design, sample composition, and analytical pipelines. As one pharmacogenomic study has previously been conducted in a Korean population^20^, we sought to evaluate the consistency of our findings with this earlier work. We compared phenotype distributions in our cohort with those reported previously. Among the ten genes analyzed, seven—including *ABCG2*, *CYP2C19*, and *CYP3A5*—showed comparable distributions (Supplementary Figure S5), whereas *SLCO1B1*, *CYP2B6*, and *UGT1A1* displayed discrepancies (Supplementary Figure S6a). We further examined haplotype distributions across the two studies to identify potential causes of these discrepancies. *SLCO1B1* showed similar haplotype distributions (Supplementary Figure S6b), whereas *CYP2B6* and *UGT1A1* displayed marked differences. These findings suggest that the observed discrepancies in phenotypes are largely attributable to differences in cohort composition and analytical approaches.

Following this comparison between the two Korean cohorts, we extended the analysis to additional populations (Figure 5, Supplementary Figure S7, and Supplementary Table S8). Compared with East Asians, although 66 of 159 gene–phenotype combinations showed statistically significant differences, the mean effect size was only 0.051, indicating minimal divergence between Koreans and other East Asians. Consistent with the haplotype-level analysis, these results suggest that pharmacogenomic phenotypes in Koreans are broadly similar to those observed in other East Asian populations.

**Figure 5.**
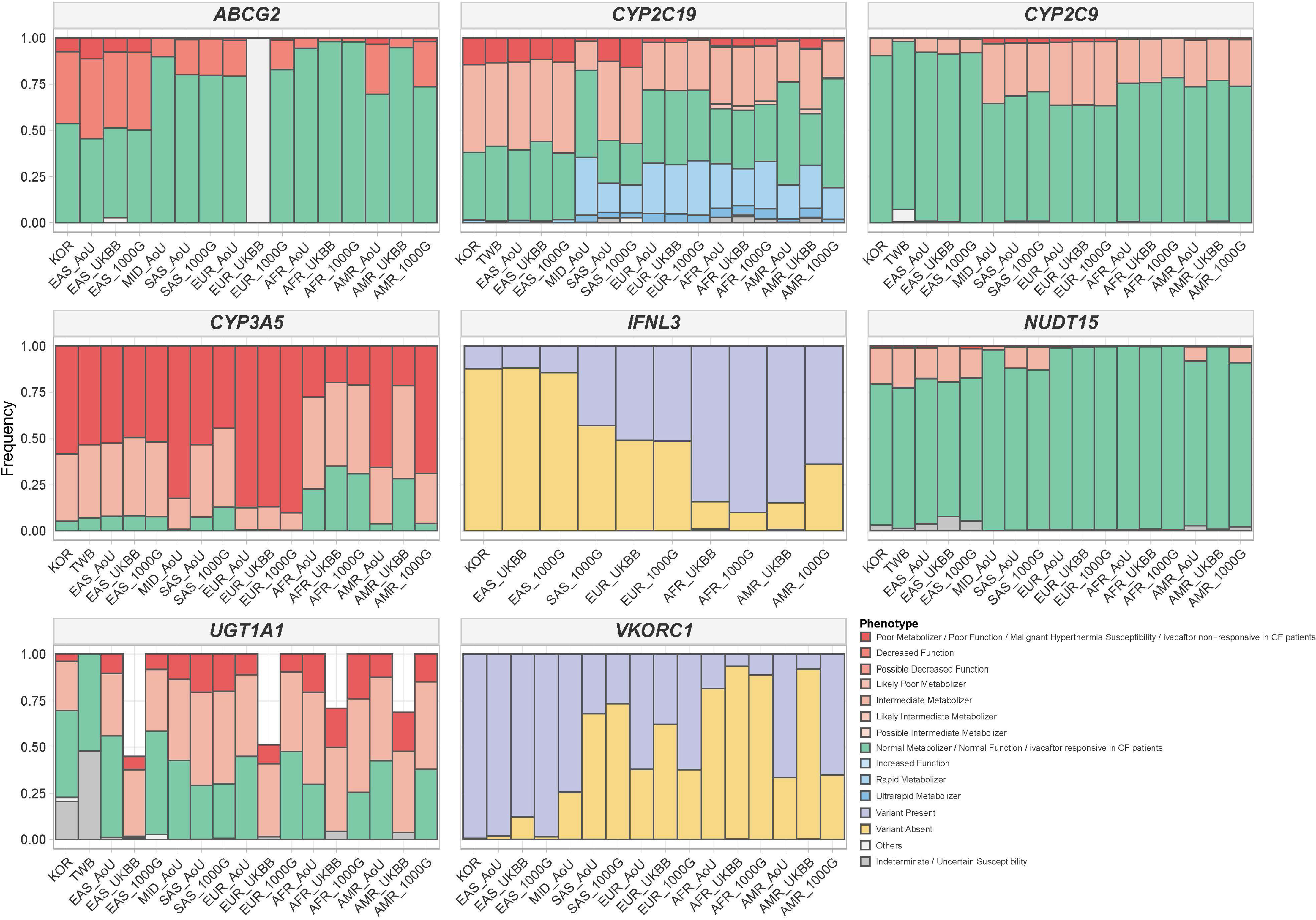
Pharmacogenomic phenotype distributions for eight genes, including *CYP2C19*, *CYP2C9*, and *VKORC1*, across different ancestry and cohort groups.

By contrast, comparisons with non–East Asian populations revealed substantial differences. For VKORC1, variant-associated phenotypes were markedly enriched in East Asians, showing a 49.934% higher prevalence relative to non–East Asian populations (effect size = 0.602). For *ABCG2*, 39.033% of Koreans carried the decreased-function phenotype compared with 12.5% in non–East Asians and only 3.189% in Africans, representing the lowest observed frequency. For *CYP2C19*, the intermediate metabolizer phenotype was more frequent in Koreans (47.405% vs 28.806%), whereas rapid metabolizers were rare in Koreans but enriched in non–East Asians (effect size = 0.262). For *CYP2C9*, Koreans had a greater proportion of normal metabolizers (90.373% vs 70.372%) and fewer intermediates (9.42% vs 27.67%), with corresponding effect sizes of 0.151 and 0.14. *UGT1A1* exhibited variability both between and within ancestry groups.

Collectively, these results demonstrate that Koreans and other East Asians share broadly similar phenotype profiles, whereas pronounced differences exist when compared with non–East Asian populations. These findings emphasize the importance of ancestry-informed prescribing, particularly for genes such as *VKORC1*, *ABCG2*, *CYP2C19*, and *CYP2C9*.

## Discussion

Although pharmacogenomics is essential for realizing the vision of precision medicine, its development has historically been constrained by technical, economic, and logistical challenges^2,39,40^. Despite these barriers, most advances in the field have originated from large-scale studies involving European populations, leaving non-European groups substantially underrepresented in pharmacogenomic research^15–17^. While progress has been made, studies involving East Asian populations remain limited in both scale and depth^19–21^. This imbalance represents a significant obstacle to the equitable implementation of genotype-guided prescribing. To help address this disparity, the present study characterized pharmacogenomic variants in a large Korean cohort of 14,490 individuals using genome-wide array data.

Our analysis revealed high frequencies of pharmacogenetically important variants in the Korean population, particularly in *CYP2C19*, *VKORC1*, and *CYP3A5* (Figure 2c and Figure 2d). For *CYP2C19* and *VKORC1*, allele distributions were consistent with those reported in other East Asian cohorts but diverged markedly from those observed in European and African populations (Figure 4d and Figure 4f). Approximately 47% of Koreans in our cohort were *CYP2C19* intermediate metabolizers, for whom CPIC guidelines recommend alternative antiplatelet therapy, and nearly 99% carried a warfarin-sensitivity *VKORC1* genotype—highlighting the importance of genotype-guided dosing. These findings have direct implications for optimizing clopidogrel and warfarin therapy^41–44^. Moreover, for *CYP3A5*, over 94% of individuals were classified as poor or intermediate metabolizers (Figure 3c), indicating markedly reduced tacrolimus clearance in this population and underscoring the clinical necessity of genotype-based dose adjustment^45^. In contrast, variants in *SLCO1B1* were frequent but predominantly associated with haplotypes of normal function (Figure 2c, Figure 2d, and Supplementary Figure S2a), illustrating that variant presence does not necessarily equate to pharmacogenomic functional impact. Collectively, these results emphasize the value of ancestry-informed prescribing and the integration of population-specific pharmacogenomic data into clinical practice guidelines.

When comparing our cohort with a previous Korean pharmacogenomic study, most genes exhibited broadly similar phenotype distributions (Supplementary Figure S5), with minor discrepancies for *SLCO1B1* and *CYP2B6* that were attributable to differences in allele definitions and annotation pipelines (Supplementary Figure S6a). By contrast, *UGT1A1* demonstrated considerable variability not only between Korean cohorts but also across global studies, where phenotype distributions differed substantially even within the same ancestry group (Supplementary Figure S6d and Figure 5). This inconsistency likely reflects technical limitations in accurately capturing variants at the complex *UGT1A1* locus^46^, including alignment challenges arising from high sequence similarity with other *UGT1A* family members and the difficulty of detecting the TA-repeat expansion that defines *UGT1A1* 28 using array- or short-read–based methods^47,48^. In addition, the accuracy of variant calling is strongly influenced by the quality of reference genomes and gene annotations^49^. These methodological constraints, together with differences in variant interpretation tools^29,50^, underscore the need for standardized pharmacogenomic annotation pipelines and advanced sequencing technologies to ensure consistent and accurate phenotype inference.

In this study, we identified not only common allele combinations but also several previously unreported haplotype structures (Supplementary Figure S1 and Supplementary Table S7). Expansion to a large-scale cohort substantially increased the number of detectable polymorphic sites (Figure 1a, Figure 1d, and Figure 2a), thereby providing the resolution necessary to uncover haplotype combinations that had not been observed previously. These findings suggest that continued scaling of population-level analyses may reveal additional Korean- or East Asian–specific variant patterns that were overlooked in smaller studies. Although these novel haplotypes were rare, their detection in multiple individuals indicates that they represent genuine variant patterns in Koreans or East Asians, consistent with findings from other Asian populations^51–54^.

For example, a *DPYD* variant combination [c.1627A>G (*5) + c.2303C>A (rs56005131)] was uniquely identified in the Korean population (Supplementary Table S7). Among these, *DPYD \**5 (rs1801159) has been reported to partially reduce enzymatic activity^55^ and was observed at a relatively high allele frequency of approximately 22% in our cohort. In addition, previous functional studies have demonstrated that c.2303C>A (rs56005131) causes a moderate reduction in dihydropyrimidine dehydrogenase (DPD) enzyme activity, retaining approximately 47–50% of normal function^56^. Therefore, the coexistence of these two variants may further impair DPD activity and potentially alter fluoropyrimidine metabolism. However, the functional and clinical consequences of this combined genotype remain uncharacterized, underscoring the need for future studies to investigate the pharmacogenetic effects of such variant combinations in East Asian populations.

Similarly, a *CYP4F2* variant combination [*3 (rs2108622) + *17 (rs3093105)] was also observed in the Korean cohort. Both alleles are associated with reduced *CYP4F2* enzymatic activity, which decreases vitamin K oxidation and may result in slightly higher warfarin dose requirements^57^. Notably, *CYP4F2* *17 (c.1414A>G, p.T472A) was recently defined by PharmVar in 2024 through long-read sequencing and haplotype-phasing analyses, representing one of the newly characterized alleles with reduced or potentially loss-of-function enzymatic activity^58^. Although each allele individually exerts only a mild effect, their coexistence could further attenuate *CYP4F2* function, warranting additional investigation to clarify its pharmacogenetic relevance in East Asians. These novel variant patterns likely account for some of the ambiguous or unassigned phenotypes observed in our analysis. At the same time, they highlight a key limitation of current annotation tools such as PharmCAT^29^ and Stargazer^50^, which rely on fixed allele catalogs. As a result, variants not represented in these catalogs—often those more common in non-European populations—can be overlooked. Consequently, East Asian individuals may appear to carry fewer pharmacogenomic variants than Europeans, not because of genuine genetic differences but due to discovery bias (Figure 4a and Figure 4b). The increasing use of long-read sequencing technologies, together with the expansion of large, ancestry-specific cohorts supported by standardized annotation pipelines, will be essential to reveal the full spectrum of pharmacogenomic diversity and to advance equitable precision medicine.

Several limitations of this study should be acknowledged. First, our analysis could not comprehensively evaluate certain genomic regions due to technical constraints. Although the genomic array included markers on sex chromosomes, imputation was not feasible because the Korean WGS reference panel lacked sex chromosome data. In addition, complex loci such as *CYP2D6*^31,59^ and *HLA*^32,60^ were excluded, as they are difficult to resolve using short-read sequencing or array-based genotyping platforms. Second, most known pharmacogenomic variants and corresponding clinical guidelines have been established primarily using data from European populations, which may not encompass variants unique to or more prevalent in the Korean population. Consequently, some potentially important population-specific variants may have been missed. These limitations are expected to be mitigated through ongoing advancements in sequencing technologies and the discovery of population-specific pharmacogenomic variants. Despite these challenges, this study represents the largest pharmacogenomic investigation conducted in the Korean population and demonstrates that genome-wide array data, when combined with a population-specific imputation strategy, can reliably identify clinically relevant variants at scale within an ancestry-relevant context. These population-level insights will directly support the development of tailored clinical guidelines, inform national healthcare policy, and promote equitable precision medicine for Korean and East Asian patients. As a next step, expanding this framework to include more than 100,000 individuals will enable improved detection of rare variants, refined haplotype and phenotype predictions, and the establishment of robust, population-specific pharmacogenomic guidelines. Ultimately, these efforts lay the groundwork for implementing ancestry-informed precision medicine in Korea and across East Asia.

## Acknowledgments

This study was supported by an intramural grant from the National Institute of Health, Republic of Korea (grant number 2024-NI-006-01).

## Ethics approval

This study was conducted in accordance with the Declaration of Helsinki and was approved by the Institutional Review Board (IRB no. 2022-02-03-P-A). All participants provided written informed consent for the use of their genomic data in research.

## Author Contributions

S.P. and M.S. designed the study. S.P and M.S wrote the manuscript. S.P. and M.S. analyzed the data. C.P., Y.K., and B.K. assisted with writing of the manuscript. All the authors have read and agreed to the current version of this manuscript.

